# Revisiting Gaza mortality estimates: adjusted for non-sampling zero-survivor households

**DOI:** 10.1101/2025.08.06.25333116

**Authors:** Ron Roberts, Joel Vos

## Abstract

Current Gaza mortality estimates systematically undercount deaths by failing to account for zero-survivor households. We argue that adjusting for this factor and multi-family cohabitation yields significantly higher figures than previously reported. If we assume 3% zero-survivor households with 75% multi-family cohabitation, mortality estimates increase to 152,395-179,555 direct and indirect deaths (6.92-8.16% of pre-war population).

## Introduction

Mortality estimates from the Gaza war beginning October 7, 2023, face systematic underestimation across all methodologies (see Table 1). The Gaza Ministry of Health (GMoH) relies primarily on hospital records, but 654 documented attacks on healthcare facilities have severely compromised record-keeping capacity.^1^ Bodies may remain unreported due to inaccessible hospitals or cultural burial practices.^2^ Airwars reports substantial undercounting in GMoH’s named casualty lists.^3^ Additionally, zero-survivor households may result in bodies remaining unrecovered, unidentified, and unreported when buried in mass graves or rubble.

**Table 1:**
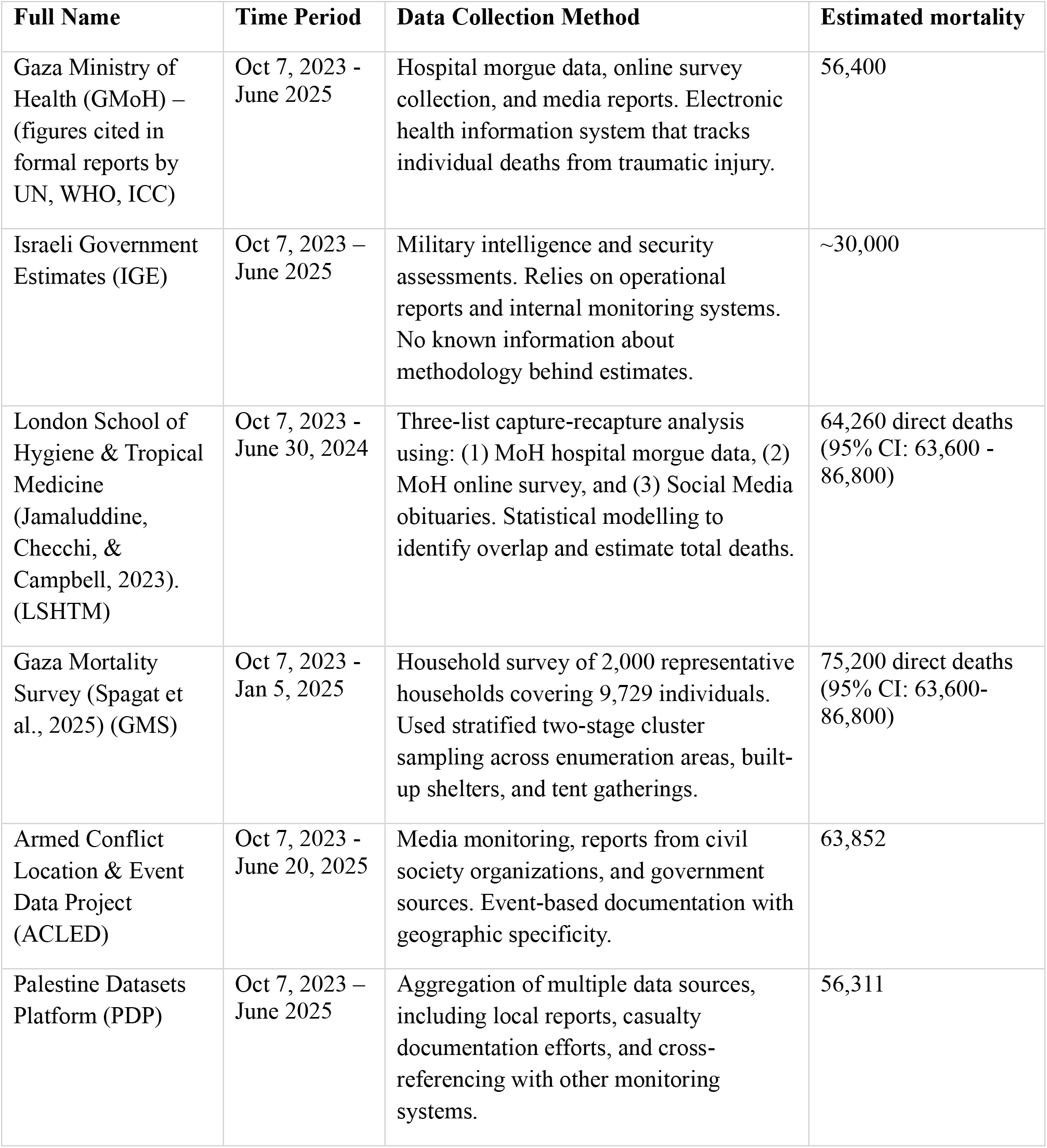
Data Collection Methods by Source.

| **Full Name** | **Time Period** | **Data Collection Method** | **Estimated mortality** |
| --- | --- | --- | --- |
| Gaza Ministry of Health (GMoH) – (figures cited in formal reports by UN, WHO, ICC) | Oct 7, 2023 - June 2025 | Hospital morgue data, online survey collection, and media reports. Electronic health information system that tracks individual deaths from traumatic injury. | 56,400 |
| Israeli Government Estimates (IGE) | Oct 7, 2023 – June 2025 | Military intelligence and security assessments. Relies on operational reports and internal monitoring systems. No known information about methodology behind estimates. | ~30,000 |
| London School of Hygiene & Tropical Medicine (Jamaluddine, Checchi, & Campbell, 2023). (LSHTM) | Oct 7, 2023 - June 30, 2024 | Three-list capture-recapture analysis using: (1) MoH hospital morgue data, (2) MoH online survey, and (3) Social Media obituaries. Statistical modelling to identify overlap and estimate total deaths. | 64,260 direct deaths (95% CI: 63,600 - 86,800) |
| Gaza Mortality Survey (Spagat et al., 2025) (GMS) | Oct 7, 2023 - Jan 5, 2025 | Household survey of 2,000 representative households covering 9,729 individuals. Used stratified two-stage cluster sampling across enumeration areas, built-up shelters, and tent gatherings. | 75,200 direct deaths (95% CI: 63,600-86,800) |
| Armed Conflict Location & Event Data Project (ACLED) | Oct 7, 2023 - June 20, 2025 | Media monitoring, reports from civil society organizations, and government sources. Event-based documentation with geographic specificity. | 63,852 |
| Palestine Datasets Platform (PDP) | Oct 7, 2023 – June 2025 | Aggregation of multiple data sources, including local reports, casualty documentation efforts, and cross-referencing with other monitoring systems. | 56,311 |

Online data collection methods face limitations from Gaza’s damaged communications infrastructure, with 70% of networks destroyed and frequent blackouts precisely when mortality reporting would occur. ^4^ The London School of Hygiene & Tropical Medicine’s capture-recapture analysis explicitly excluded missing persons potentially deceased under rubble. ^5^

The Gaza Mortality Survey (GMS) by Spagat et al. attempted to overcome these limitations through face-to-face interviews yet acknowledged “downward bias” from non-sampling households with zero survivors or no surviving adults. ^6^ This critical bias remains unaddressed in all current estimates.

Gaza’s exceptionally high population density (6,024 people/km^2^ average, reaching 161,532 people/km^2^ in Beach Camp) dramatically increases vulnerability compared to other conflict zones (20-300 people/km^2^ in Rwanda, Syria, Bosnia, Darfur, Iraq, and Yemen). ^7^ This density exacerbates full-family casualties from single strikes. The UN reports 35% of Gaza buildings destroyed by February 2024, suggesting substantial numbers remain buried. ^8^

Additionally, multi-family cohabitation—prevalent at 50-75% pre-war and likely higher post-displacement^9^—further complicates mortality accounting. The documented 2,200 zero-survivor households as of May 2025^10^ likely underestimates the true figure and doesn’t account for the 1.41 multiplier for individuals missing under rubble. ^11^

Table 1 outlines major mortality estimation methodologies. Table 2 presents our adjusted confidence intervals incorporating zero-survivor households and multi-family cohabitation levels applied to the Gaza Household Survey figures^6^, with equivalent findings to the capture-recapture study based on morgue records, online surveys, and social media reports^5^. While our data assume the same ratio of direct to indirect deaths as the Gaza Mortality Survey, it is probable that there remains a significant undercount of indirect deaths, for instance due to starvation, non-communicable diseases, endemic infectious disease, epidemics and neonatal health complications^12,13,14^; these may be higher due to the unique population density and household composition in Gaza, which may lead to a quicker spread of diseases and increased vulnerability of humanitarian transport routes in densely populated areas.

**Table.2:** Adjusted Confidence Intervals by Household Loss x Multi-Family Cohabitation (95% CI)

|  | **Multi-Family Cohabitation** |  |  |  |
| --- | --- | --- | --- | --- |
| **% Zero-survivor households** | 0% MFC | 25% MFC | 50% MFC | 75% MFC |
| 0.5% (1,900) | 77,455 - 104,610 | 79,033 – 106,188 | 80,610 – 107,765 | 82,188 - 110,920 |
| 1.0% (3,800) | 86,760 - 113,915 | 89,915 – 117,070 | 93,070 – 120,230 | 99,380 - 126,540 |
| 2.0% (7,600) | 105,370- 132,525 | 111,680 – 138,835 | 118,000 – 145, 160 | 124,310- 151,470 |
| 3.0% (11,400) | 123,980- 151,135 | 133,445 - 160,600 | 142,930 – 170,090 | 152,395- 179,555 |
| 4.0% (15,200) | 142,590- 169,745 | 155,210 - 182,365 | 167,830 – 194,985 | 180,450- 207,605 |
| 5.0% (19,000) | 161,200- 188,355 | 176,975 - 204,130 | 192,790 – 219,950 | 208,565- 235,725 |

Each cell shows the adjusted 95% confidence interval for total deaths, assuming systematic exclusion of zero-survivor households. Formula: Adjusted CI = Base CI + Additional Deaths, where additional deaths = Homes $\times [(1 - \text{MF}\%) + \text{MF}\% \times 2] \times 4.9$

Mortality figures during conflicts typically undergo substantial upward revision post-conflict (by factors of 0.7-23.3). ^15,16,17,18^Given Gaza’s unprecedented population density and documented destruction scale, current estimates therefore likely represent significant undercounts.

While it is impossible to make a precise estimation of the multiplier factors for zero-survivor households and indirect deaths, our calculations indicate the importance of adjusting the figures for the non-sampling of zero-survivor households and household composition. As the authors of the Gaza Mortality Survey noted in their conclusions, the ratio of non-violent deaths to violent deaths may well have grown since the period of data collection. Therefore, our own figures are still likely to be conservative and to seriously under-estimate the present death toll. Accurately recording the complete magnitude of destruction serves dual purposes: preserving historical truth while honouring all victims’ suffering and fulfilling essential legal obligations under international humanitarian law.

## Data Availability

All data produced in the present work are contained in the manuscript

https://www.medrxiv.org/content/10.1101/2025.06.19.25329797v3

## References

1. World Health Organization. Health Cluster Bulletin: Gaza Strip. Regular situation reports citing Gaza MoH data, 2023-2025.

2. Stamatopoulou-Robbins S. The Human Toll: Indirect Deaths from War in Gaza and the West Bank, October 7, 2023 Forward. Watson Institute for International and Public Affairs. The Human Toll: Indirect Deaths from War in Gaza and the West Bank, October 7, 2023 Forward | Costs of War.

3. Airwars. Gaza Casualty Assessment: Methodological Review of Gaza Ministry of Health Figures. Airwars Report, May 2025.

4. United Nations Office for the Coordination of Humanitarian Affairs (OCHA). Humanitarian Situation Update #297 | Gaza Strip. June 18, 2025. https://www.ochaopt.org/content/humanitarian-situation-update-297-gaza-strip;

5. Jamaluddine Z, Abukmail H, Aly S, Campbell OMR, Checchi F. Traumatic injury mortality in the Gaza Strip from Oct 7, 2023, to June 30, 2024: a capture-recapture analysis. The Lancet. 2025;405(10477):469–477. doi: 10.1016/S0140-6736(24)02678-3.

6. Spagat M, Guha-Sapir D, Abu-Raddad LJ, et al. The Gaza Mortality Survey: A population-based assessment of mortality in the Gaza Strip, October 7, 2023 - January 5, 2025. medRxiv 2025; doi: 10.1101/2025.06.19.25329797 [Preprint].

7. Palestinian Central Bureau of Statistics (PCBS). Population Density Report: Gaza Strip 2023. PCBS Statistical Yearbook, 2023.

8. United Nations Satellite Centre (UNOSAT). Damage assessment of the Gaza Strip. February 2024.

9. United Nations Relief and Works Agency (UNRWA). Occupied Palestinian territory. 2015 Emergency Appeal. Annual Report. 2015. https://www.unrwa.org/sites/default/files/content/resources/2015_opt_emergency_appeal_annual_report.pdf

10. New Arab. ‘They want to erase us’: Israel wipes out over 2,200 Palestinian families in Gaza. May 26, 2025. https://www.newarab.com/news/israel-wipes-out-over-2200-palestinian-families-gaza

11. UN Office of the High Commissioner for Human Rights (OHCHR). Update Report on the Situation in Gaza. May 6, 2024. https://www.ohchr.org/sites/default/files/documents/countries/opt/20241106-Gaza-Update-Report-OPT.pdf

12. Khatib R, McKee M, Yusuf S. Counting the dead in Gaza: difficult but essential. The Lancet. 2024;404(10449):237–238.

13. Sah S. Infectious diseases are being allowed to run rampant in Gaza. BMJ 2024; 387:q2186.

14. Jamaluddine Z, Chen Z, Abukmail H, et al. Crisis in Gaza: scenario-based health impact projections. Report one: Feb 7 to Aug 6, 2024. https://gaza-projections.org/gaza_projections_report.pdf x(accessed Oct 10, 2024).

15. Hagopian A, Flaxman AD, Takaro TK, et al. Mortality in Iraq associated with the 2003-2011 war and occupation: findings from a national cluster sample survey by the University Collaborative Iraq Mortality Study. PLoS Med. 2013;10(10):e1001533.

16. Verpoorten, M. ‘How many died in Rwanda?’, Journal of Genocide Research, 2020, 22(1), pp. 94–114.

17. World Bank, The Toll of War: Economic and Social Consequences of the Conflict in Syria, 2022, Washington, DC. World Bank.

18. United Nations Development Programme (UNDP) Assessing the Impact of War on Development in Yemen, 2021. New York. UNDP.

